## Supplementary material for "Self-Supervised Data-Driven Approach Defines Pathological High-Frequency Oscillations in Human": Suppl. data

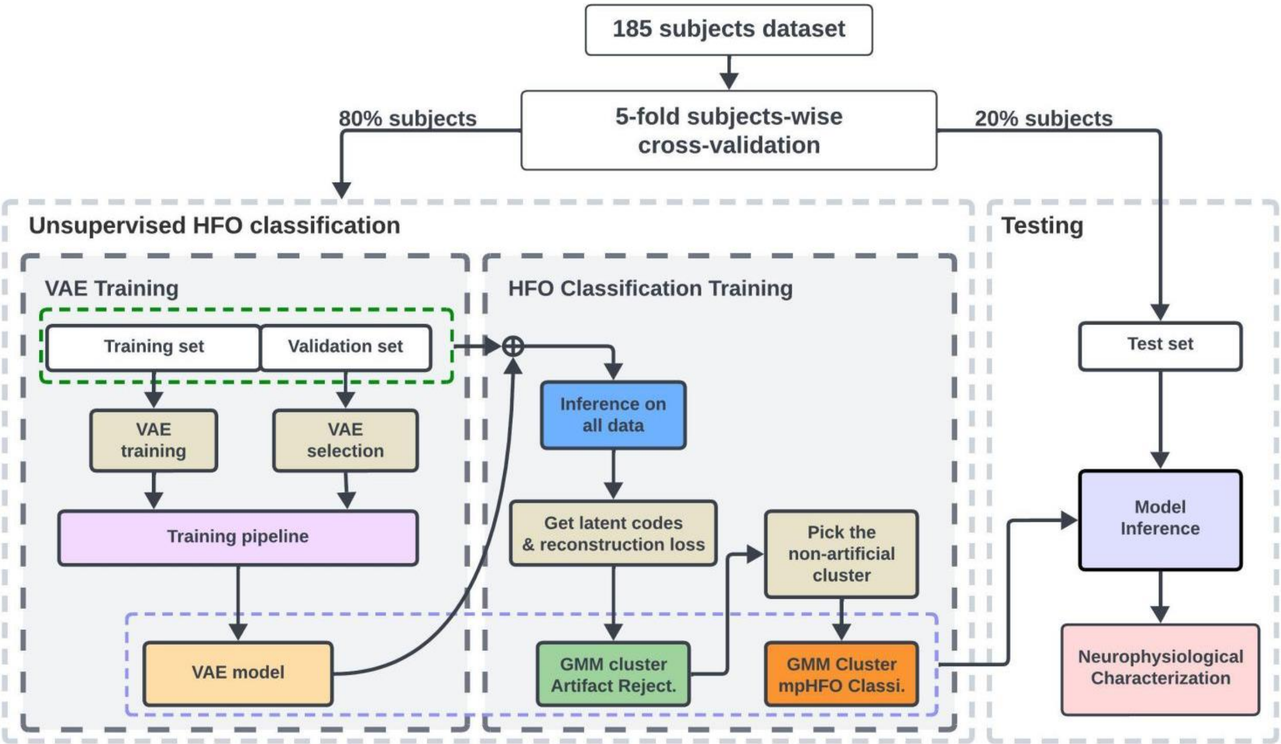


**Supplementary Figure 1. VAE training and HFO classification workflow.** The VAE was validated using subject-wise five-fold cross-validation. In each fold, a VAE model was trained and tested on a specific test set, with 20% of the subjects uniformly selected from three datasets to form the test set and the remaining 80% used as the training/validation set. During VAE training, ten subjects per dataset were randomly designated as the validation set, which was not used for direct training the network but instead served to monitor model performance and prevent overfitting during the training. The remaining data was used as the training set. The VAE was trained for 80 epochs, and the model with the lowest validation loss, recorded after each epoch, was selected as the "best model" for this fold. In the HFO classification stage, the trained VAE model was used to infer latent codes (mean values) and calculate reconstruction loss for each HFO event (time-frequency plot) in both the training and validation sets. These features were then clustered using a hierarchical two-stage Gaussian Mixture Model (GMM) pipeline. In the first stage, a GMM with $k = 2$ was fitted, and we picked the cluster with a higher reconstruction loss cluster as the "artificial" cluster (mArtifact) and a lower reconstruction loss cluster as the "non-artificial" cluster, Using reconstruction loss as artifact detection is well recognized in the abnormal detection domain using VAE. The non-artificial cluster was further divided by a second GMM to separate mpHFO and non-mpHFO clusters based on the assumption that mpHFO and non-mpHFO exhibit fundamental morphological differences. We used minimal clinical information to assign a pathological (mpHFO) label and physiological (non-mpHFO) label for the resulting cluster, i.e., the cluster with a higher resection percentage in seizure-free patients was deemed pathological. After the training of VAE and GMMs was completed, the VAE encoder and both GMMs were capable of assigning classification labels to HFOs in the test set, characterizing both mpHFO and non-mpHFO categories. The VAE and GMM training were conducted in each fold so that all of the subjects were tested once across five folds.

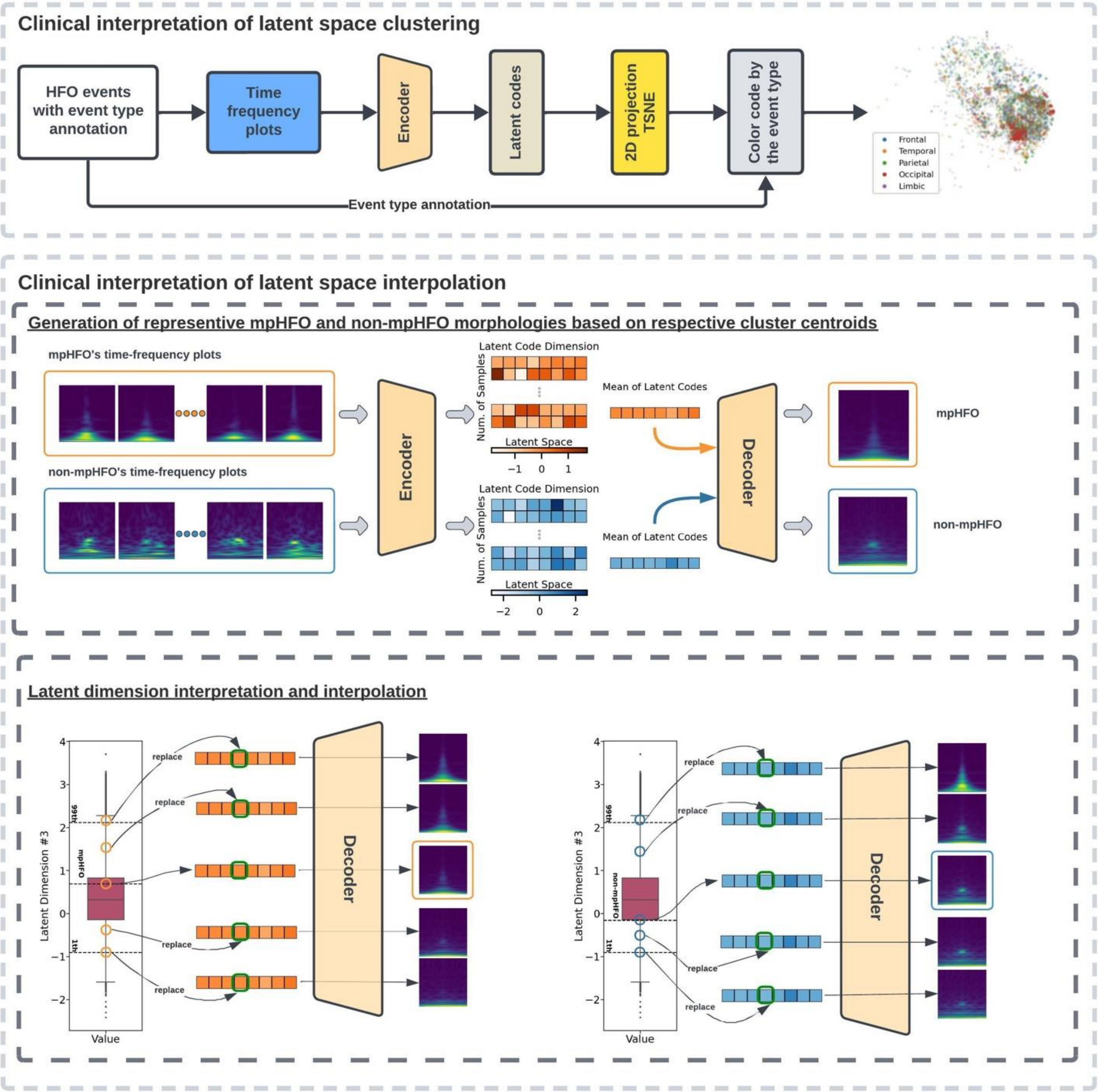


**Supplementary Figure 2. Model Interpretation Analysis Workflow.** *Clinical interpretation of latent space clustering:* The latent codes generated from time-frequency representations of HFOs were visualized by projecting them into a two-dimensional space, allowing us to observe the distribution of different HFO subcategories (e.g., anatomical locations). *Generation of representative mpHFO and non-mpHFO morphologies based on respective cluster centroids:* The VAE latent space was further interpreted through the VAE decoder, which generated meaningful time-frequency plots from latent codes. A classic approach to understanding the VAE and clustering performance involved generating the centroid for each cluster. The centroid for both mpHFO and non-mpHFO clusters was calculated by averaging the latent codes of each predicted group, producing representative latent codes for mpHFO and non-mpHFO clusters (orange and blue vectors). The decoder then generated time-frequency plots from these centroids, revealing that mpHFO and non-mpHFO have distinctive frequency-band characteristics, with mpHFO having higher intensities in the HFO and gamma bands. *Latent*

*dimension interpretation and interpolation*: To explore the neurophysiological significance of individual latent dimensions, we applied dimension-specific perturbations. Left: Starting with the mean latent code for mpHFO (orange, middle dashed line in the boxplot), we linearly interpolated values along a specific dimension (Dimension No. 4, highlighted in the green box in this figure) from the 99th percentile (top dashed line in the boxplot) down to the mean, and from the mean to the 1st percentile (bottom dashed line in the boxplot), holding all other dimensions constant. Each perturbation generated a new latent code where only the value in Dimension No. 4 changed. We used the decoder to transform these perturbed latent codes into new time-frequency plots, which allowed us to interpret how variations in Dimension No. 4 influenced HFO characteristics. Right: The same perturbation process was applied to non-mpHFO (blue) to determine whether the effects in this dimension were specific to mpHFO or non-mpHFO. Please note that, for population-level analysis, we performed a continuous interpolation from the 1st to the 99th percentile within values in this dimension.

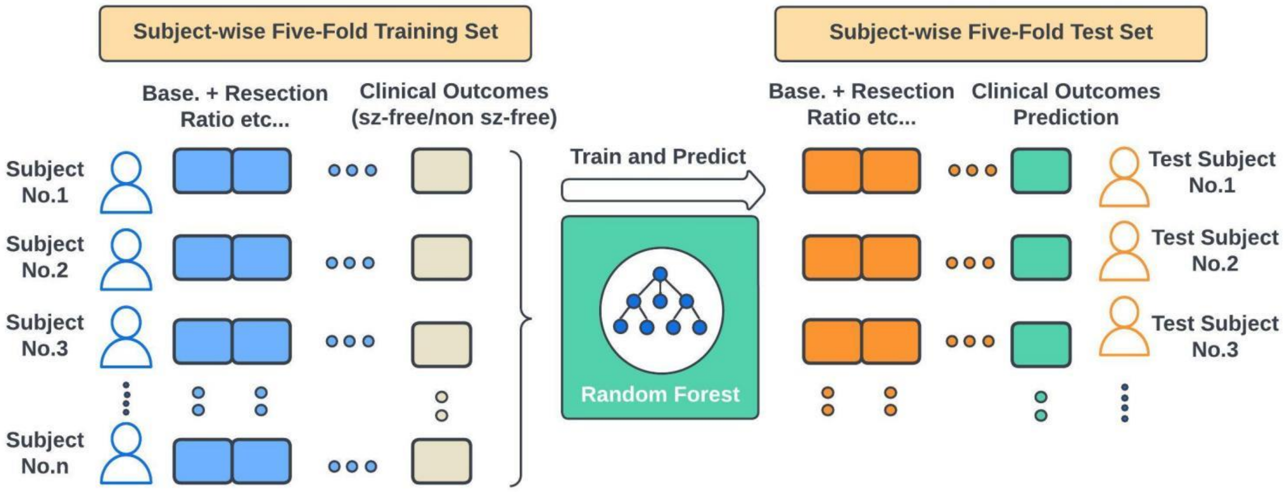


**Supplementary Figure 3. Building postoperative seizure outcome prediction model using HFO information.** We used a random forest model to conduct forward prediction, evaluating the predictive power of mpHFO classification and comparing it with other HFO classification methods and clinical standards (baseline demographic information and SOZ). The goal was to predict whether a patient achieved postoperative seizure freedom. As our model was trained using subject-wise five-fold cross-validation, we obtained five VAE + GMM models through this setup. In each fold, we utilized subjects from the training and validation sets to train the random forest and then predicted postoperative seizure freedom for subjects in the test set. This setup ensured that the entire pipeline—VAE + GMMs + random forest—remained uninfluenced by any data from the test set, providing a robust assessment of the framework's generalization ability. Given the class imbalance in surgical outcomes (approximately 70% of patients achieved seizure freedom), we reported the F1 score in the main text, as it more effectively reflects model performance on imbalanced datasets.

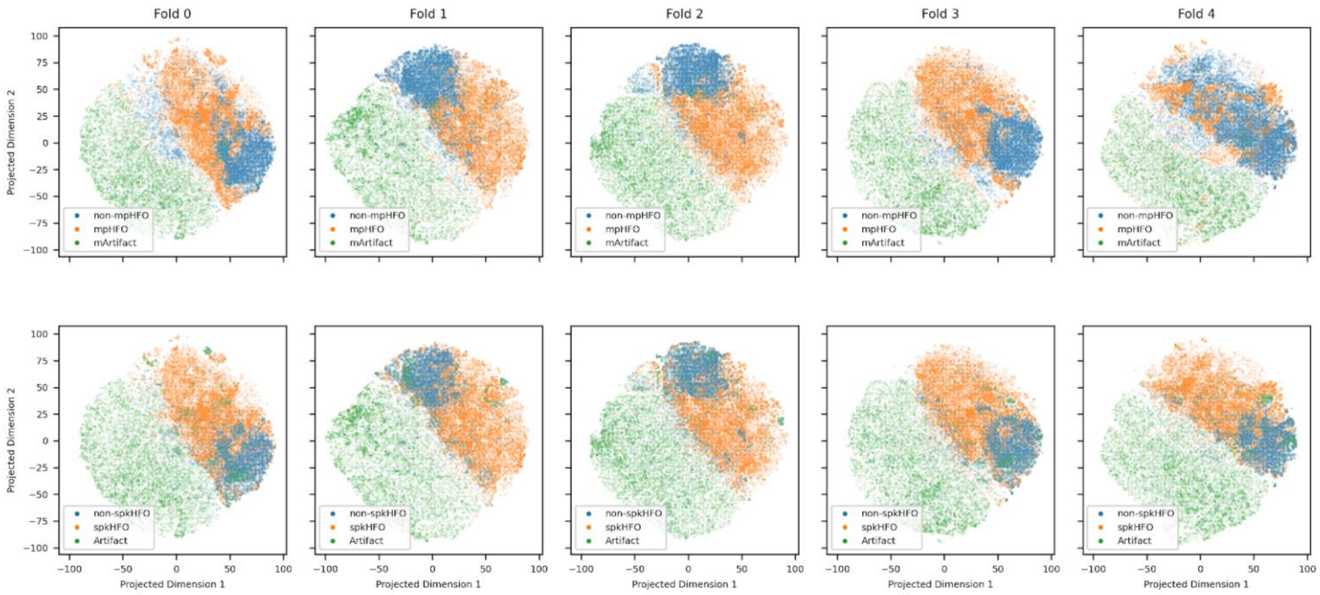


**Supplementary Figure 4. Latent space comparison:** This figure illustrates the 2D projection of the latent space, color-coded according to classifications from a VAE (mpHFO, non-mpHFO, and mArtifact) and from conventional supervised methods (spkHFO, non-spkHFO, and Artifact) from PyHFO, evaluated across five folds. It highlights the alignment between VAE predictions and conventional supervised classifications, demonstrating that classifiers developed from unsupervised learning can achieve performance comparable to those developed from supervised methods.

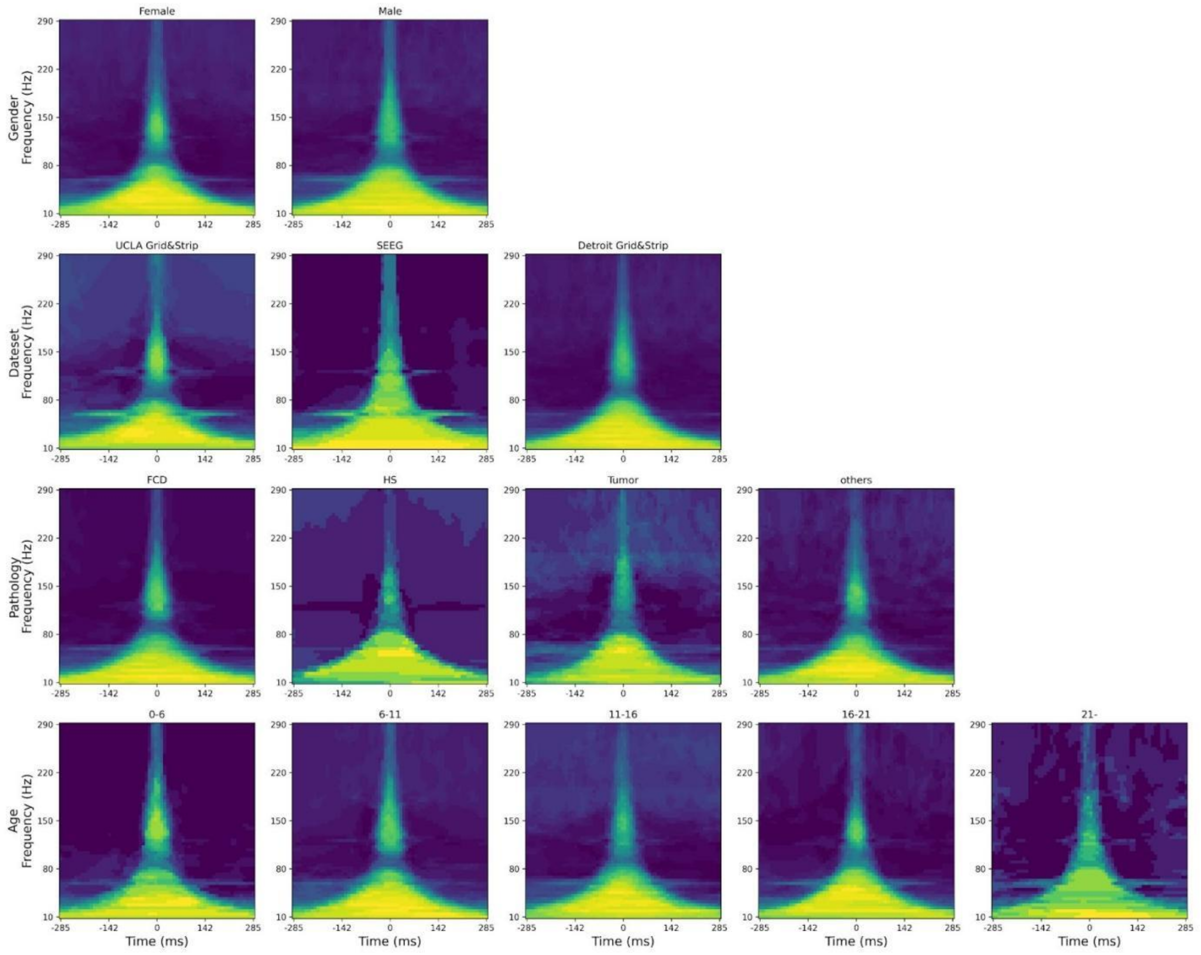


**Supplementary Figure 5. mpHFO shares similar morphology across the variables.** The morphological analysis of mpHFOs' time-frequency plot is shown based on subgroups, each corresponding to a specific variable. Consistent morphological features of mpHFOs are seen across the subgroups. mpHFOs have higher values throughout the HFO band (> 80 Hz), around the center point (0 ms, where HFOs were detected) than non-mpHFOs; furthermore, higher values of mpHFOs at the sub-HFO band (10-80 Hz) throughout the time window compared to non-mpHFOs are seen.

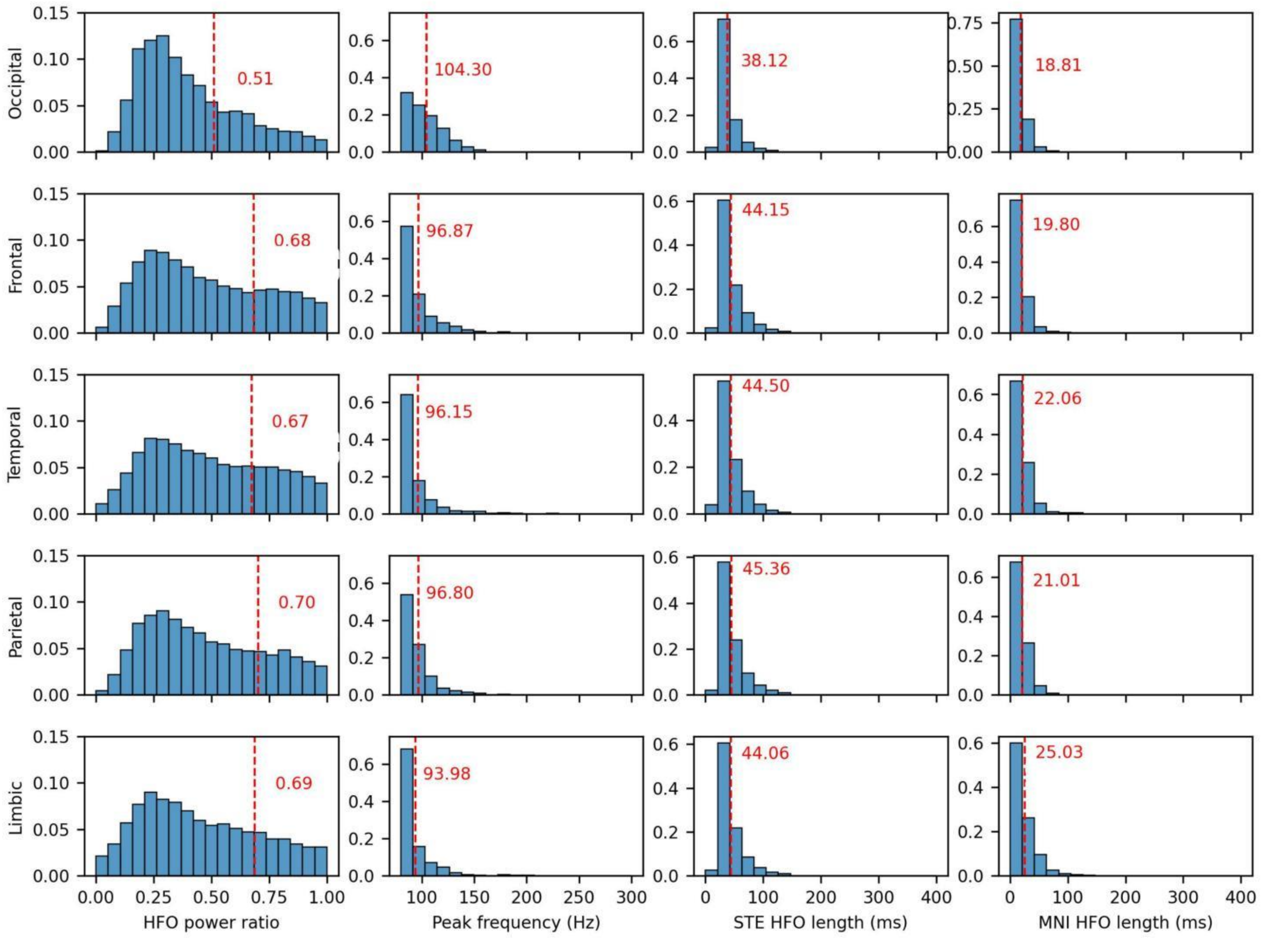


**Supplementary Figure 6. Conventional physiological non-mpHFO characteristics across different anatomical locations.** This figure displays multiple aspects of HFOs: the HFO power ratio in the first column, HFO peak frequency in the second column, STE HFO length in the third column, and MNI HFO length in the fourth column. The HFO power ratio is defined as the ratio of the signal power within ±50ms of the HFO center to the power within the entire time window of ±285ms. The peak frequency is identified as the frequency component that has the most power within the 80-300 Hz range. Both STE and MNI HFO lengths are measured in milliseconds as the duration of the HFO event. Each histogram's mean value is annotated within its respective panel. Additionally, statistical comparisons revealed that physiological HFOs in the occipital region exhibited lower power ($p$<0.001, one-sided t-test), higher peak frequency ($p$<0.001, one-sided t-test), and shorter durations in both STE and MNI HFOs ($p$<0.001 for all, except in the frontal location where $p$<0.01, one-sided t-test) compared to those in other anatomical locations.

.

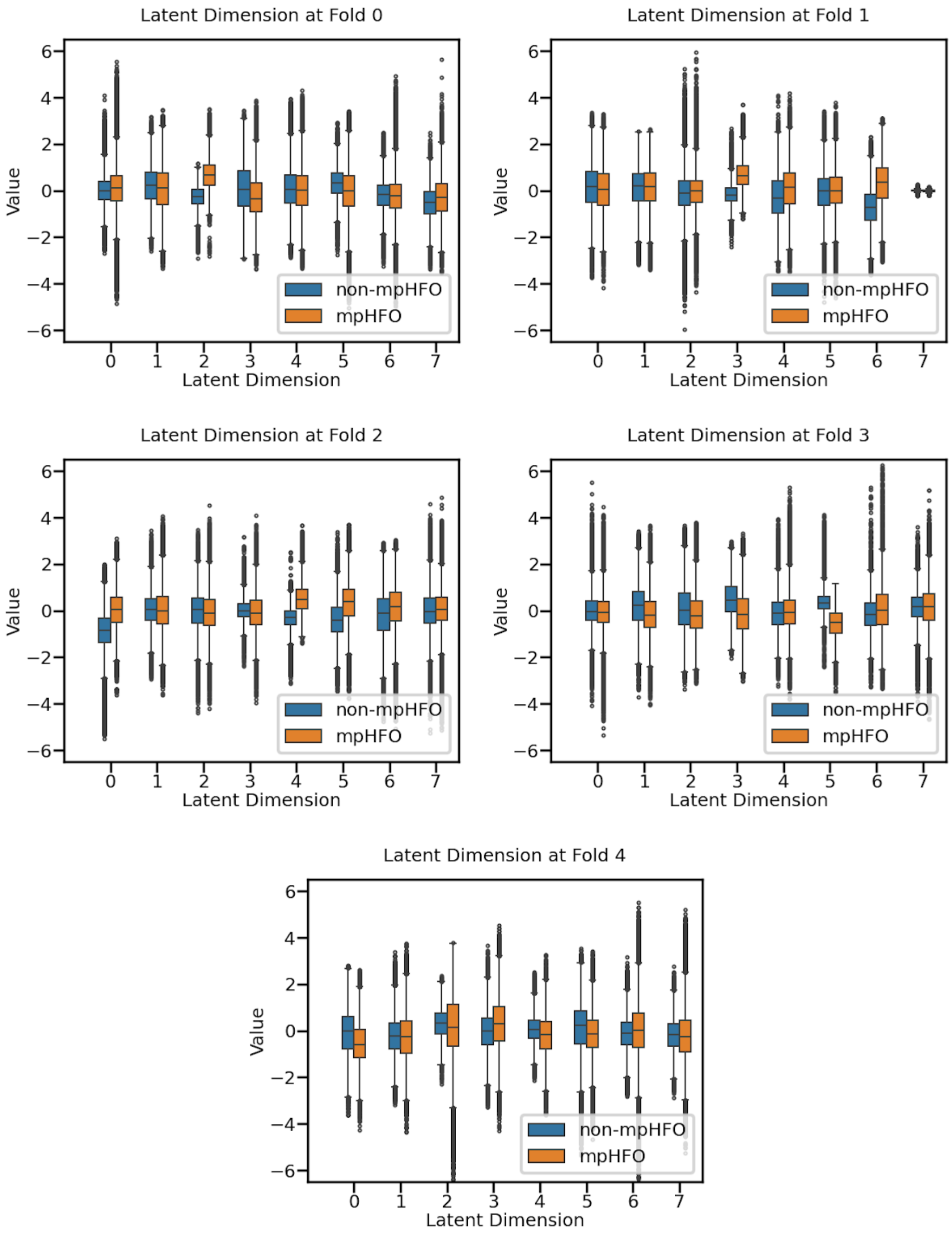


**Supplementary Figure 7. Full latent space visualization across five folds.** This visualization presents the distribution of each latent dimension, trained within five folds and depicted by separating mpHFO (in orange) and non-mpHFO (in blue). It is evident that mpHFO and non-mpHFO exhibit significant separation in some dimensions while others show less distinction.

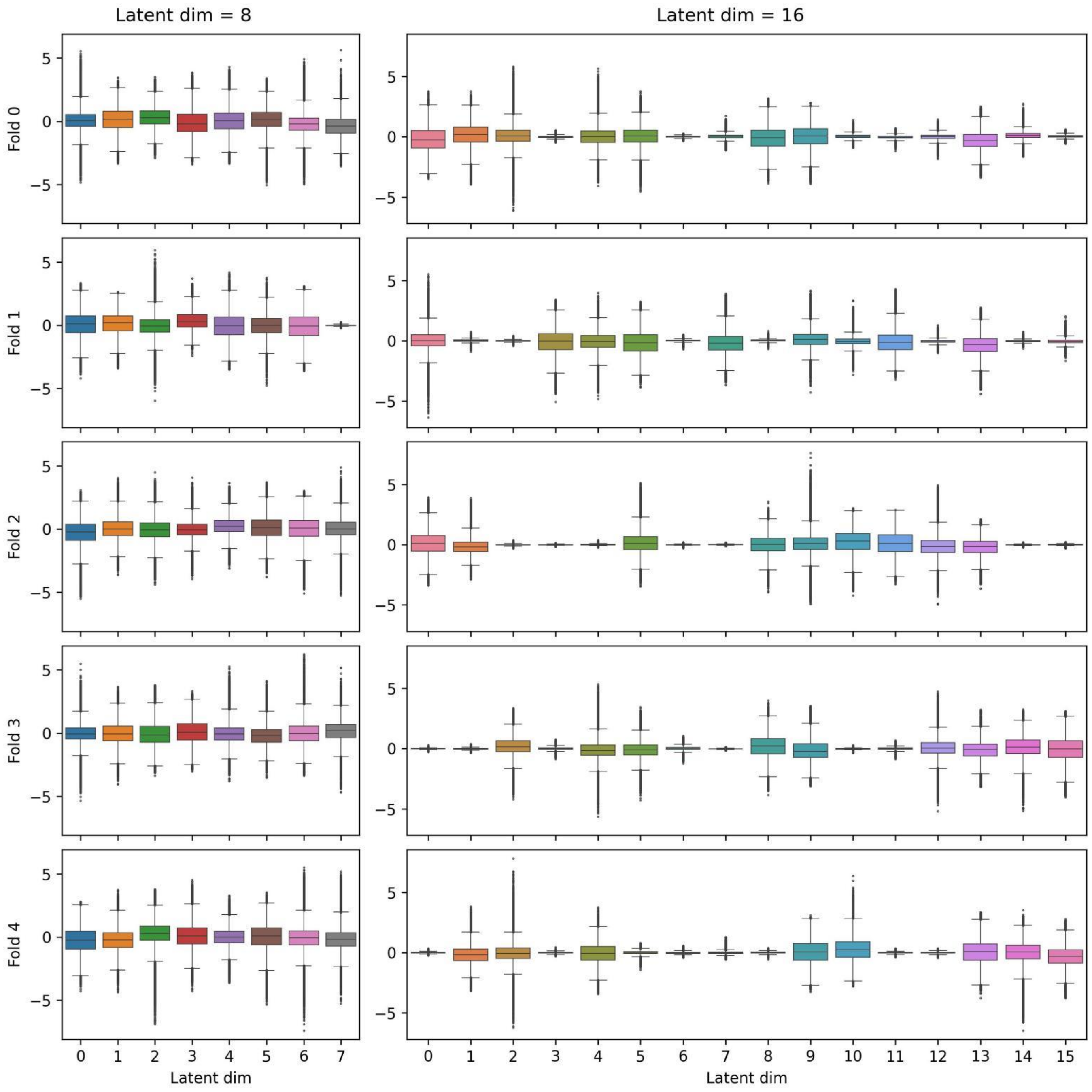


**Supplementary Figure 8. Ablation study on different latent dimensions.** The left panel shows the distribution of dimensions for a VAE model trained with a latent dimension of 8, across five folds. The right panel displays the distribution for the same model configuration but with a latent dimension of 16. Several dimensions in the 16-dimension model do not spread well, indicating that these dimensions are underutilized during training. This compactness suggests a potential reduction in dimensionality. Based on these observations and the more effective utilization of dimensions seen in the boxplot for the 8-dimension model, we chose to proceed with a latent dimension of 8, as it demonstrates optimal use of all available dimensions during training.

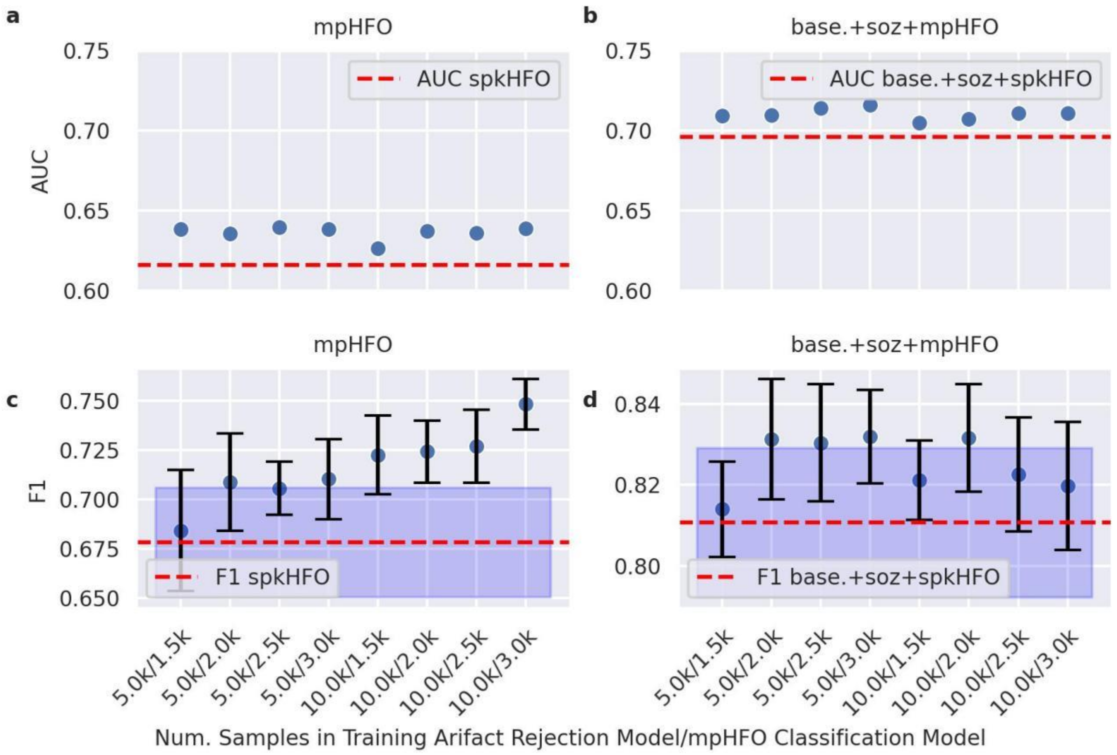


**Supplementary Figure 9. Ablation study on predicting surgical outcomes using different sample sizes for training GMM models.** We conducted experiments using different stratified sampling to train GMM models and investigated how capping the number of samples per subject used to train the artifact rejection and mpHFO classification impacts surgical outcomes prediction performance metrics such as AUC and F1 score. We compared models capped at 10,000 or 5,000 samples per subject for artifact rejection and 3,000, 2,500, 2,000 or 1,500 samples for mpHFO classification. (a) Comparison of AUC scores for mpHFO models trained from different stratified sampling sizes against those from spkHFO models. (b) A similar comparison for base.+soz+mpHFO (baseline demographic, soz resection, resection ratio of mpHFO) against base.+soz+spkHFO (baseline demographic, soz resection, resection ratio of spkHFO). (c) Comparison of F1 scores for mpHFO models trained from different stratified sampling sizes against those from spkHFO models. (d) Similar procedure as in (c), Similar comparison for base.+soz+mpHFO against base.+soz+spkHFO.

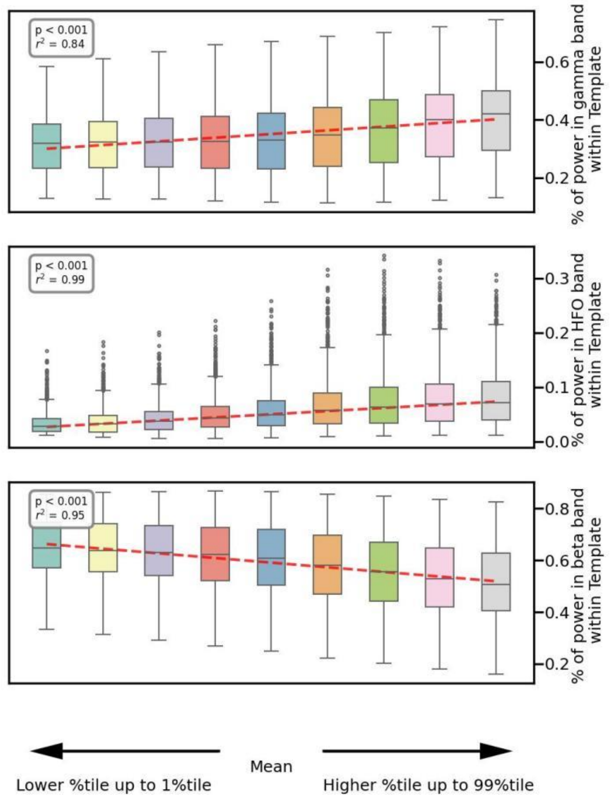


**Supplementary Figure 10. Power percentages in different frequency bands (gamma, HFO, and beta) within a cone-shaped template region: Top panel:** the percentage of power in the gamma band. A statistically significant positive correlation is observed ($p < 0.001$, $r^2 = 0.84$), indicating an increase in gamma power as the latent dimension is adjusted. **Middle panel:** the percentage of power in the HFO band. A strong positive correlation is evident ($p < 0.001$, $r^2 = 0.99$), suggesting a pronounced increase in HFO power with changes in the latent dimension. **Bottom panel:** the percentage of power in the beta band. A significant negative correlation is present ($p < 0.001$, $r^2 = 0.95$), indicating a decrease in beta power as the latent dimension varies. In all panels, the x-axis represents a range from lower percentiles (up to the 1st percentile) to higher percentiles (up to the 99th percentile), with the mean value marked at the center. The red dashed lines indicate the regression trends for each frequency band.

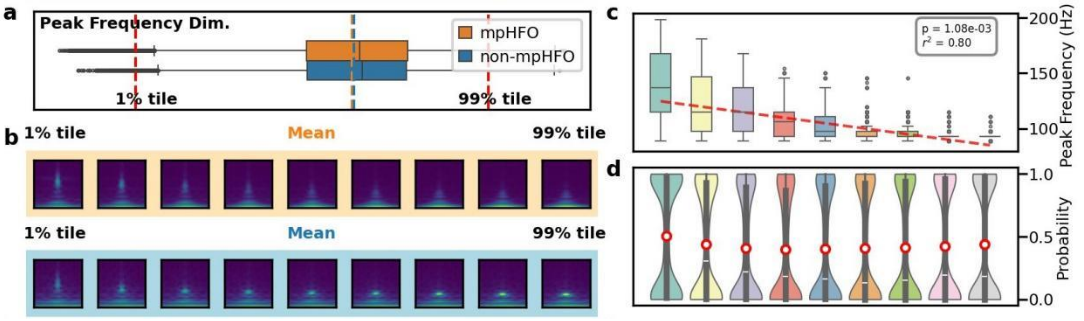


**Supplementary Figure 11. Peak frequency dimension in the latent space.** (a) Peak frequency dimensional visualization. A similar visualization of another dimension is shown. This latent space represents the peak frequency of HFOs. (b) The output of the decoder traversing the dimension, displayed in the image sequence, showed a descending trend in peak frequency from upper to lower percentiles of the value of that dimension. (c) At the population level, the box plots indicated a negative correlation between the peak frequency dimension value and the peak frequency in decoded images, with a trend line fitted from the median of each box. (d) Distribution of model probability scores for each sample. The red circles indicate the mean probability scores, showing the average confidence of perturbed events was around 0.5 (unchanged).

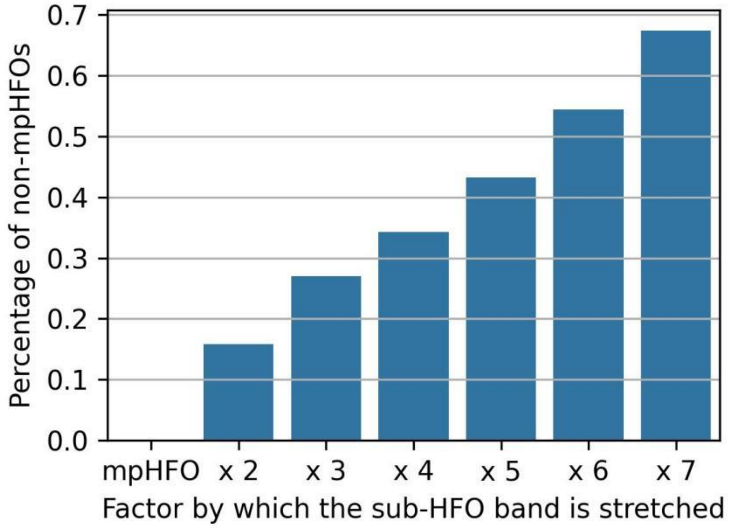


**Supplementary Figure 12.** Effect of stretching the sub-HFO band on mpHFOs. The bar plot shows the percentage of mpHFO events that were transformed into non-mpHFOs at varying levels of time-domain perturbation. The x-axis indicates the factor by which the sub-HFO band was slowed down, ranging from the original signal (labeled 'mpHFO') to factors of 2 through 7. The y-axis represents the percentage of events classified as mpHFOs by the model after each perturbation. As the slowing factor increases, a larger proportion of events transition from mpHFO to non-mpHFO, suggesting that the gamma-band activity plays a crucial role in the model's mpHFO classification.

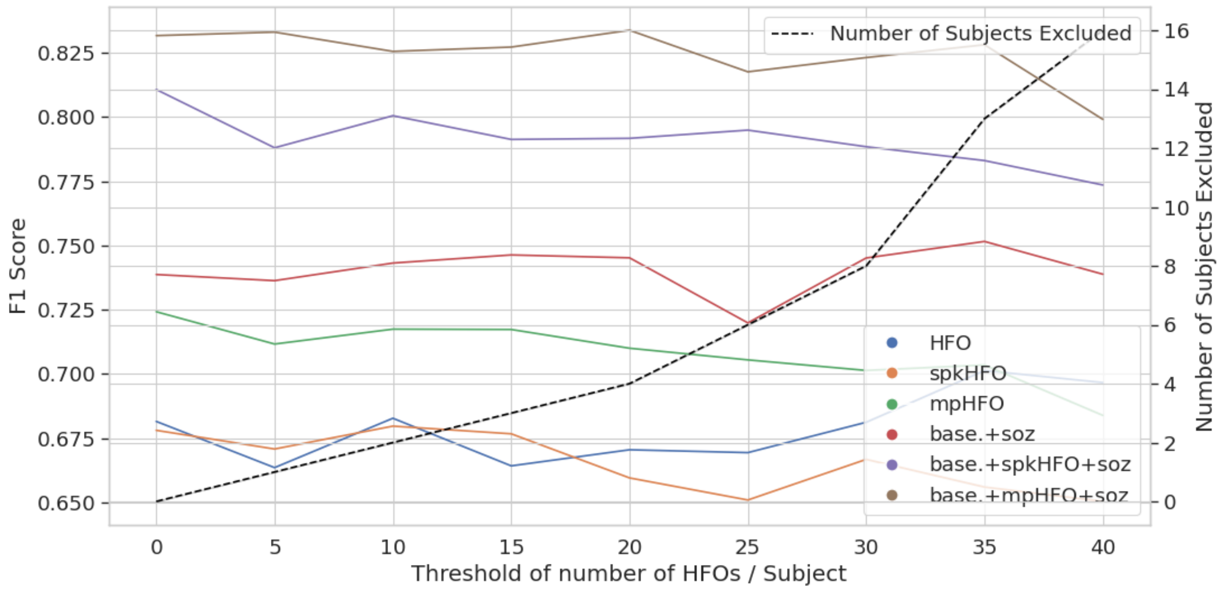


**Supplementary Figure 13. Ablation study on predicting surgical outcomes by excluding patients with fewer HFOs:** We conducted an ablation study to assess the impact of excluding patients with a smaller number of high-frequency oscillations (HFOs) on the performance of our Random Forest model in predicting post-surgical outcomes. In addition to the results presented in the main manuscript, which included all patients, we compared the F1 scores for predicting post-surgical outcomes by selectively including patients with a minimum of 10, 20, 30, and 40 HFOs. The F1 scores were plotted for different feature sets used in constructing the Random Forest model. The results indicate that the F1 score remains consistent even when including only patients with more than 40 HFOs, demonstrating the robustness of our self-supervised framework.

**Supplementary Table 1**

| Dataset | No. Event | No. Artifact | No. spkHFO | No. non-spkHFO |
|---|---|---|---|---|
| Detroit grid/strip | 35610 | 4374 | 20481 | 10755 |
| UCLA grid/strip | 103360 | 14578 | 54455 | 34327 |
| UCLA SEEG | 35861 | 7886 | 21748 | 6227 |
| All STE HFOs | 174831 | 26838 | 96684 | 51309 |
| Detroit grid/strip | 36279 | 6494 | 26953 | 2832 |
| UCLA grid/strip | 124064 | 42848 | 69266 | 11950 |
| UCLA SEEG | 351236 | 275550 | 50434 | 25252 |
| All MNI HFOs | 511579 | 324892 | 146653 | 40034 |

**Supplementary Table 2**

| **Feature (s)** | **Accuracy** | **F1** |
|---|---|---|
| HFO | 0.555 (0.083) | 0.682 (0.075) |
| spkHFO | 0.568 (0.144) | 0.678 (0.139) |
| mpHFO | 0.626 (0.078) | 0.724 (0.080) |
| base.+ soz | 0.652 (0.062) | 0.739 (0.049) |
| base.+ soz + spkHFO | 0.739 (0.099) | 0.811 (0.092) |
| base.+ soz + mpHFO | 0.752 (0.084) | 0.832 (0.066) |

The mean accuracy, F1 scores, and standard error of the mean for the random forest models trained on the training subjects using five-fold cross-validation and evaluated on the test subjects across five folds with different features are presented. In the main manuscript, we focused on F1 scores due to the class imbalance in the postoperative surgical outcome labels (with 69.8% of subjects achieving seizure freedom).

**Supplementary Table 3**

| | STE |
|---|---|
| Filter freq (Hz) | [80, 300] |
| RMS window (s) | 0.003 |
| Min window (s) | 0.006 |
| Min gap (s) | 0.01 |
| Min osc (count) | 6 |
| RMS thres (in SD) | 5 |
| Peak thres (in SD) | 3 |
| Epoch len (ms) | 600 |

| | MNI |
|---|---|
| Filter freq (Hz) | [80, 300] |
| Epo CHF (s) | 60 |
| Per CHF (%) | 0.95 |
| Min win (s) | 0.01 |
| Min gap (s) | 0.01 |
| Thrd perc (%) | 0.999999 |
| Base seg (s) | 0.125 |
| Base shift (0-1) | 0.5 |
| Base thrd (0-1) | 0.67 |
| Base min (%) | 5 |
| Epoch time (ms) | 10 |
| Seed | 0 |

**Supplementary Table 4:**

| Dataset | No. Event | STE HFOs only | MNI HFOs only | Exactly same | Overlap |
|---|---|---|---|---|---|
| Detroit grid/strip | 71889 | 20873 | 21542 | 540 | 14191 |
| UCLA grid/strip | 227424 | 90169 | 69465 | 810 | 33085 |
| UCLA SEEG | 387097 | 22943 | 338318 | 146 | 12772 |
| Sum | 686410 | 133985 | 429325 | 1496 | 60048 |

We calculated the statistics of the number of overlapped HFO events between two detectors. During the training, there were at most only around 8.96% ( [60048+1496]/686410) events double counted (despite the stratified sampling). Furthermore, please note that the overlapped events do not necessarily mean the STE and MNI represent the same event, but we tried to merge them in calculating the resection ratio of events in clinical validation.

### Supplementary Methods

#### Interpretability analysis:

*Time-frequency plot characteristics of pathological and physiological HFOs:* We examined whether the time-frequency plots (scalograms) of mpHFOs identified by the VAE differ from non-mpHFOs using a t-test on each pixel similar as outlined in preceding studies (Ref#9-10 of the main manuscript). The null hypothesis was that scalogram values for mpHFOs are the same or lower than for non-mpHFOs. We binarized p-values less than 0.05 and created a characteristics map that highlights mpHFOs' distinct morphology in both frequency and time domains. To examine differences in the time-frequency scalograms (64 × 64 pixels) of mpHFOs identified by the VAE versus non-mpHFOs, we compared pixel-wise intensity values across all subjects, following a methodology similar to previous studies. For each pixel (x, y), we created two datasets: $S_{mpHFO}(x, y)$, containing the intensity values f(x, y) for all mpHFOs, and $S_{non\text{-}mpHFO}(x, y)$, containing the intensity values f(x, y) for all non-mpHFOs. We performed one-tailed t-tests to determine if $S_{mpHFO}(x, y)$, was significantly greater than $S_{non\text{-}mpHFO}(x, y)$, testing the null hypothesis that scalogram values for mpHFOs were the same as or lower than those for non-mpHFOs. If a pixel's p-value was less than 0.05, we set the pixel to 1 in the resulting characteristics map, indicating significant morphological differences in mpHFOs at that location; otherwise, we set it to 0. This binarized characteristics map highlights morphological distinctions in mpHFOs across time and frequency domains, enhancing our understanding of the VAE's classification criteria for pathological HFOs.

*Latent space 2D visualization:* To visualize the latent space learned by the VAE, we extracted latent codes from the time-frequency plots of each event. We applied t-distributed Stochastic Neighbor Embedding (t-SNE) using the cuML package,[55] to project these latent codes into two-dimensional space (**Supplementary Figure 2: Clinical interpretation of latent space clustering**). For better visualization, we randomly sampled up to 200 data points per subject in plotting the 2D latent space. Each point was color-coded based on different classifications of HFOs— defined either morphologically by VAE (mpHFO/non-mpHFO/mArtifact) or through prior knowledge (spkHFO/non-spkHFO/artifact). Additionally, we visualized the latent space using subject demographic subcategories such as sex, recording site/type, age, pathology, and anatomical locations. To further explore the significance of these factors, we employed a statistical test to evaluate the relationship between HFO morphology and demographic information, as well as the anatomical location events.

*Statistical tests to evaluate the dependence of HFO morphology on the significance of different demographic subject-level information and anatomic locations of the HFO events variability*

We evaluated whether HFO morphology depends on demographic factors. If HFO morphology is dependent on a particular variable (for example, the subject's sex), then the latent codes of HFOs labeled with the corresponding subcategories (female or male) should be distinguishable. Consequently, a classifier designed to identify true labels from the latent codes of HFOs would achieve higher accuracy compared to a classifier trained with randomly assigned labels.

This methodology can be illustrated using sex as an example. For each fold of the cross-validation, we randomly selected three subjects from each subcategory (male, n=3; female, n=3). We randomly sampled 100 HFO events from each subcategory (100 HFO events for male subjects and 100 HFO events for female subjects). This resulted in 200 latent codes (n_subcategories * 100) for training a logistic regression classifier. For computing accuracy, we repeated the sampling process for the left-out subjects in each fold, resulting in one accuracy data point. We repeated this sampling, training, and testing process five times (trials) for each fold. Thus, we obtained 25 trials in total (5 folds * 5 trials) for a classifier determining sex based on actual sex labels.

Next, we designed a surrogate classifier by randomizing the labels. For each fold and each trial, the same latent codes sampled for the true-label case were used, but the sex labels were randomized. Specifically, we randomly shuffled the subjects and assigned the label "male" to the first three and "female" to the last three. A surrogate logistic classifier was then trained using these randomized sex labels. For testing, we used the same data samples as in the true-label case but randomized the sex labels again. This process yielded another 25 accuracy samples for the surrogate classifier. A one-tailed t-test was employed to assess whether the true-label classifier's accuracy was significantly higher than that of the surrogate.

We conducted two similar tests to evaluate whether HFO morphology depends on anatomical factors. One was to assess whether the morphology of presumed physiological HFOs (from preserved regions in seizure-free patients) depended on anatomical locations, and the second was to evaluate the dependence of presumed pathological HFO morphology (from SOZ regions) on anatomical locations. Each EEG channel in each subject was assigned a unique anatomical location. For each fold and each trial, we sampled an equal number of HFO events (n=100, totaling 500) from each anatomical location across all patients for a given HFO type. A multi-class logistic regression model was trained, and the confusion matrix was computed based on the 500 samples from the test subject in the same fold. This process was repeated five times for each fold. Thus, for each HFO type, we obtained 25 confusion matrices, and the average confusion matrix and overall accuracy were computed. For the surrogate classifier, label randomization was done at the channel level: all samples from a particular channel that were initially labeled as "Frontal," for example, were reassigned to the same randomly selected label, such as "Occipital." This resulted in 25 surrogate accuracy samples. A one-tailed t-test was again employed to assess whether the true-label classifier's accuracy was significantly higher than that of the surrogate.

*Latent space disentanglement on latent dimension perturbation:*

We interpolated along specific dimensions of the latent space in VAE to understand their significance in relation to neurophysiological characteristics (**Supplementary Figure 2: Clinical interpretation of latent space interpolation**). To determine the dimensions that contributed the most to the classification decision boundary, we plotted the distribution of the values in the latent codes conditioned on mpHFO and non-mpHFO for each latent dimension. Dimensions in which the distribution conditioned on mpHFOs was significantly distinct from that conditioned on non-mpHFOs were deemed pathological. Since each latent code represents a time-frequency plot, we interpreted the morphological features each dimension represents by sweeping through these dimensions and analyzing changes in resulting images. Specifically, we focused on the morphological impact on two representative latent codes, which are the mean of the latent vectors of all identified mpHFO and non-mpHFO events. For each latent dimension, we interpolated values from the 1st to the 99th percentiles (excluding outliers) while keeping other dimensions constant, then characterized the resulting images decoded from VAE decoder. We searched for three types of latent dimensions: 1) a pathological dimension, along which time-frequency plots transition from mpHFOs to non-mpHFOs; 2) a beta band power dimension, where beta band power in the time-frequency plots increases through interpolation; and (3) a peak frequency dimension, where the peak frequency of the HFO increases through interpolation. To analyze these dimensions at a population level, we randomly selected 500 high-confidence mpHFOs ($P_{mpHFO} > 0.99$) and 500 high-confidence non-mpHFOs ($P_{mpHFO} < 0.01$), and interpolated the latent codes of these 1,000 samples on these dimensions. To quantitatively evaluate morphological changes in the resulting images, we designed metrics based on image processing techniques on the time-frequency plots as proxies for the morphological characteristics of the HFO events. To evaluate the resulting images in pathological and beta band dimensions, we calculated the ratio of the power of the interested band in the characteristic template to all of the power in the template. For the peak-frequency dimension, we averaged the time-frequency plot from –90 ms to 90 ms, constructed a frequency-power vector, and found the frequency with maximum power within 80 Hz to 290 Hz. Finally, we used the inference pipeline (encoder and GMM) to classify each generated event (time-frequency plot) and visualize the model confidence.

*Time-domain perturbation:*

We perturbed the time-domain signal of HFOs to further investigate the role of gamma-band activity. Specifically, for one mpHFO event, we applied a 4th-order Butterworth bandpass filter to decompose the signal into two components: the sub-HFO band (<80 Hz) and the ripple band (>80 Hz). We then stretched the waveform in the sub-HFO band by interpolating values between samples in the original waveform. Afterward, the ripple band signal (HFO component) was added back to the time-stretched sub-HFO signal to reconstruct the event. A time-frequency plot was generated from this combined signal to serve as input for the inference pipeline, and the model's confidence ($P_{mpHFO}$) was recorded. At the population level,  same time-domain perturbation was applied to predicted mpHFO events from subjects in the test set. We visualized the change in the model's confidence by analyzing the proportion of mpHFO events that were transformed into non-mpHFOs using a bar plot.
